# Estimating excess mortality in people with cancer and multimorbidity in the COVID-19 emergency

**DOI:** 10.1101/2020.05.27.20083287

**Authors:** Alvina G. Lai, Laura Pasea, Amitava Banerjee, Spiros Denaxas, Michail Katsoulis, Wai Hoong Chang, Bryan Williams, Deenan Pillay, Mahdad Noursadeghi, David Linch, Derralynn Hughes, Martin D. Forster, Clare Turnbull, Natalie K. Fitzpatrick, Kathryn Boyd, Graham R. Foster, DATA-CAN, Matt Cooper, Monica Jones, Kathy Pritchard-Jones, Richard Sullivan, Geoff Hall, Charlie Davie, Mark Lawler, Harry Hemingway

## Abstract

Cancer and multiple non-cancer conditions are considered by the Centers for Disease Control and Prevention (CDC) as high risk conditions in the COVID-19 emergency. Professional societies have recommended changes in cancer service provision to minimize COVID-19 risks to cancer patients and health care workers. However, we do not know the extent to which cancer patients, in whom multi-morbidity is common, may be at higher overall risk of mortality as a net result of multiple factors including COVID-19 infection, changes in health services, and socioeconomic factors.

**Methods:** We report multi-center, weekly cancer diagnostic referrals and chemotherapy treatments until April 2020 in England and Northern Ireland. We analyzed population-based health records from 3,862,012 adults in England to estimate 1-year mortality in 24 cancer sites and 15 non-cancer comorbidity clusters (40 conditions) recognized by CDC as high-risk. We estimated overall (direct and indirect) effects of COVID-19 emergency on mortality under different Relative Impact of the Emergency (RIE) and different Proportions of the population Affected by the Emergency (PAE). We applied the same model to the US, using Surveillance, Epidemiology, and End Results (SEER) program data.

**Results:** Weekly data until April 2020 demonstrate significant falls in admissions for chemotherapy (45-66% reduction) and urgent referrals for early cancer diagnosis (70-89% reduction), compared to pre-emergency levels. Under conservative assumptions of the emergency affecting only people with newly diagnosed cancer (incident cases) at COVID-19 PAE of 40%, and an RIE of 1.5, the model estimated 6,270 excess deaths at 1 year in England and 33,890 excess deaths in the US. In England, the proportion of patients with incident cancer with ≥1 comorbidity was 65.2%. The number of comorbidities was strongly associated with cancer mortality risk. Across a range of model assumptions, and across incident and prevalent cancer cases, 78% of excess deaths occur in cancer patients with Harry ≥1 comorbidity.

**Conclusion:** We provide the first estimates of potential excess mortality among people with cancer and multimorbidity due to the COVID-19 emergency and demonstrate dramatic changes in cancer services. To better inform prioritization of cancer care and guide policy change, there is an urgent need for weekly data on cause-specific excess mortality, cancer diagnosis and treatment provision and better intelligence on the use of effective treatments for comorbidities.

## Introduction

The excess risk of death in people living with cancer during the COVID-19 emergency may be due not only to COVID-19 infection, but also to the unintended health consequences of changes in health service provision, the physical or psychological effects of social distancing, and economic upheaval. Recent national evidence suggests that there is an excess of deaths, both in those infected with SARS-CoV-2, and in those with no infection(1). Optimal cancer care must balance the need to protect patients from COVID-19 infection, with continued access to early diagnosis and potentially curative treatment(2, 3). Professional associations in the US(4–6), UK(7–9) and Europe(9) have recommended revising cancer care activities for triaging of systemic anti-cancer treatment, surgery and risk-adapted radiotherapy(10). All elective surgery has been postponed in the UK(8, 9). In the US, more than a quarter of patients with cancer reported a delay in their cancer treatment in April 2020 because of COVID-19(4). Despite these recommendations for changes in healthcare services, there is a lack of near real-time data quantifying the extent of disruption due to reconfiguration in service delivery for cancer patients.

Existing publications on cancer and COVID-19 have been limited to small case series(11, 12). The US Centers for Disease Control and Prevention (CDC) and Public Health England (PHE) have identified patients with specific malignant and non-malignant conditions as at greater risk of developing severe illness from COVID-19 exposure(13–15). Patients with cancer commonly have other conditions considered to further increase their COVID-19 risk; multimorbidity in cancer is an increasing clinical concern(16, 17). Oncologists, internists and family physicians need to balance the requirement to encourage people to socially isolate, with their need to access hospital services to ensure the most effective cancer care. There is a lack of evidence on pan-cancer estimation of mortality risks according to type and number of multimorbid conditions.

Our objectives were to: 1) quantify changes in cancer care activities in near real-time from weekly multi-center hospital data; 2) estimate the prevalence, incidence and background (pre-COVID-19) 1-year mortality risk across 24 site-specific cancers; 3) estimate the prevalence of 15 CDC- and PHE-relevant co-occurring condition clusters (multimorbidity) by cancer site and their association with excess mortality and 4) estimate overall excess deaths due to the COVID-19 pandemic over a 1-year period, based on different Proportion of the population Affected by the Emergency (PAE) and Relative Impact of the Emergency (RIE), using a previously published model(18).

## Methods

### Weekly information on cancer care

Through DATA-CAN, the UK National Health Data Research Hub for Cancer(19), we obtained weekly returns for urgent cancer referrals for early diagnosis and chemotherapy attendances (from 2018 to most current data) from hospitals in Leeds, London and Northern Ireland.

### Patient cohort

We used population-based electronic health records in England from primary care data linked to the Office for National Statistics(ONS) death registration, using the open-access CALIBER resource(20, 21). The study population consisted of 3,862,012 adults aged ≥30 years, registered with a general practice from 1 January 1997 to 1 January 2017 with at least one year of follow-up data.

### Electronic health record phenotype

definitions of diseases and risk factors relevant to COVID-19 are available at https://caliberresearch.org/portal and have previously been validated(22–25). Further details are in supplementary methods. In brief, phenotypes are based on hospital and primary care information recorded in primary care, using the Read clinical terminology(version 2). We defined non-fatal, incident and prevalent cases of cancer across 24 primary cancer sites according to previously validated CALIBER electronic health record phenotypes, which included: biliary tract, bladder, bone, brain, breast, cervix, colon-rectum, Hodgkin’s lymphoma, kidney, leukemia, liver, lung, melanoma, multiple myeloma, non-Hodgkin’s lymphoma, esophagus, oropharynx, ovary, pancreas, prostate, stomach, testis, thyroid and uterus(26). We defined cancers as prevalent (diagnosed at any time prior to baseline) and incident if they occurred during follow-up. We defined 15 comorbid conditions or condition clusters involving 40 individual conditions defined by the Centers for Disease Control (CDC) or Public Health England (PHE) as associated with poor outcomes caused by severe COVID-19 infection.

### Analyses

For full details of the analyses, see supplementary methods. Briefly, we estimated incidence rates per 100,000 person years and 1-year mortality in our study population. Estimates were used to calculate the excess deaths in cancer alone and cancer plus comorbidities, based on three levels of PAE (10%, 40% and 80%) and four RIE scenarios associated with COVID-19(1.2, 1.5, 2.0 and 3.0) (additional details on plausible choice of PAE and RIE are presented in the discussion). We presented all results in Figures 2, 5, S4 and S8, but in order to represent a plausible, likely conservative, scenario for the current emergency, we chose to highlight data for a PAE of 40% and a RIE of 1.5, with a range of 1.2–2 in the results text section of the manuscript. We applied our model to US population using publicly available data from the US from the Surveillance, Epidemiology, and End Results (SEER) program on incidence and 1 year mortality (taken as 1 minus the reported survival(27)). We considered mortality data from SEER for individuals aged 40-64, 65-74 and 75+.

**Fig. 1.**
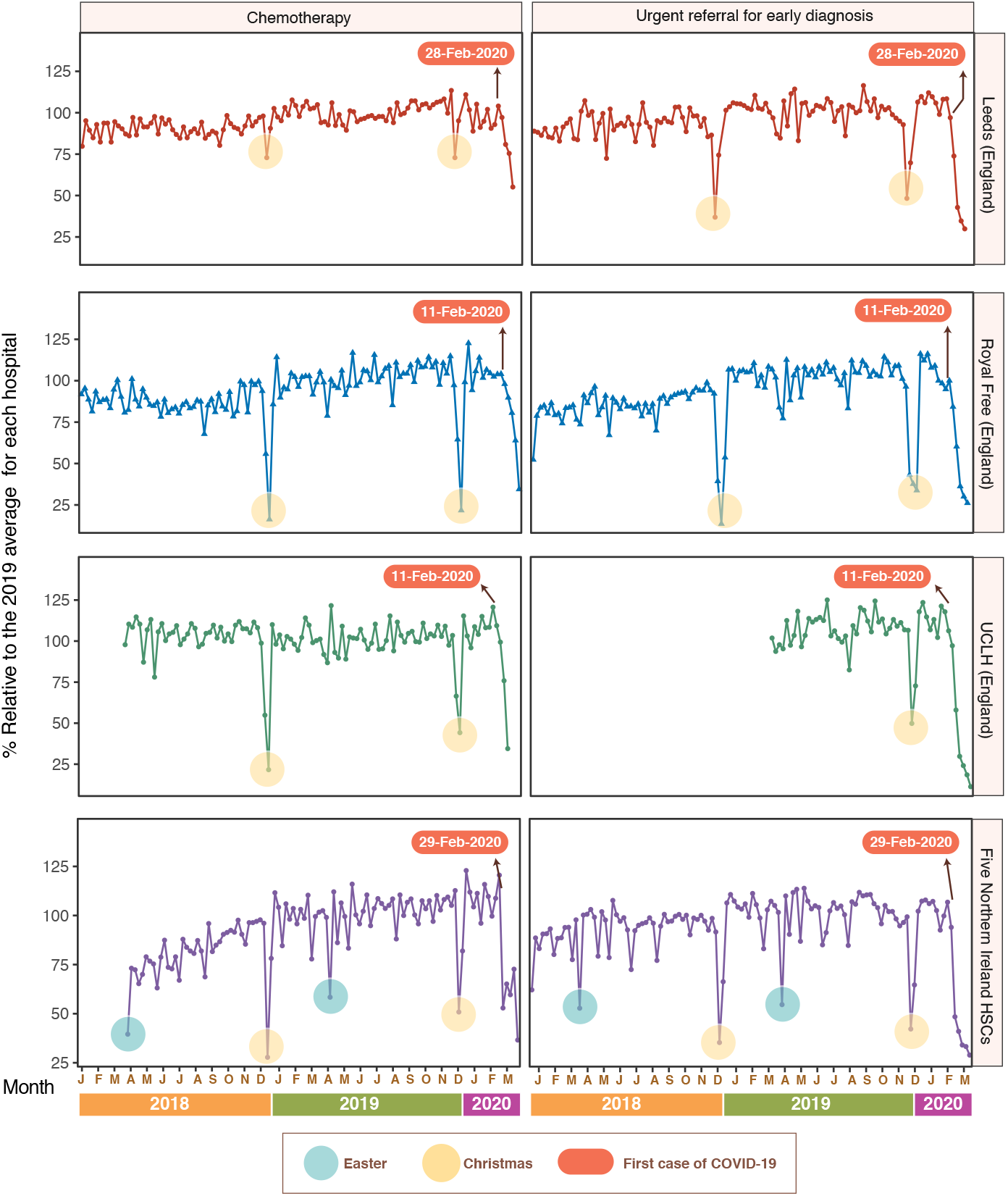
Weekly hospital data on changes in urgent referrals and chemotherapy clinic attendance from eight hospitals in the UK. The data for Northern Ireland includes five Health and Social Care Trusts (HSCs) that cover all health service provision in Northern Ireland: Belfast HSC, Northern HSC, South Eastern HSC, Southern HSC and Western HSC.

## Results

### Real-time weekly hospital data for urgent cancer referrals and chemotherapy attendances in England and Northern Ireland

We observed that a majority of patients with cancer or suspected cancer are not accessing healthcare services. Major declines in chemotherapy attendances (45%, 66%, 66% and 63% reduction; average=60%) and urgent cancer referrals for early diagnosis (70%, 74%, 89% and 71% reduction; average=76%) were observed in eight hospitals across the UK: England (Leeds Teaching Hospitals NHS Trust, Royal Free Hospital and University College London Hospitals) and Northern Ireland (all five Health and Social Care Trusts) respectively, compared to pre-emergency levels (Figure 1). These data signal the need to identify patient-specific risk factors (cancer and multimorbidity) to facilitate prioritization of service provision during the pandemic.

### Prevalence, incidence and background (pre-COVID-19) 1-year mortality for 24 cancers

The overall prevalence of any of the 24 cancers was 3.1% (117,978/3,862,012) in CALIBER (England), (Table S1). Prevalence of the 24 cancers by age, sex and year is shown in Figure S1. The age-adjusted incidence rates of the 24 cancers estimated in CALIBER was 635 per 100 000 (compared with estimates from International Agency for Research on Cancer (IARC, UK) of 590 per 100 000) (Figure S2A). The 1-year mortality across 24 cancer sites was, as anticipated, higher among incident cases of cancer, compared to prevalent cases (Figure S3).

### Estimates of excess COVID-19-related deaths by cancer site in England

When considering incident cancers in England, at COVID-19 PAE of 40%, we estimated 2,509, 6270 and 12,543 excess deaths at RIE of 1.2, 1.5, and 2 respectively (Figure 2A). Higher numbers were seen among prevalent cancers than incident cancers when estimating absolute excess deaths, since the number of prevalent cases were higher than the number of incident cases observed over 1 year, e.g., for PAE 40%, we observed 11,645 excess deaths (range 4,655–23,287) at RIE of 1.5 (range 1.2–2) (Figure S4). When considering both prevalent and incident cancers together at COVID-19 PAE of 40%, we estimated 17,915 excess deaths (range 7,164–35,830) at RIE of 1.5 (range 1.2–2).

**Fig. 2.**
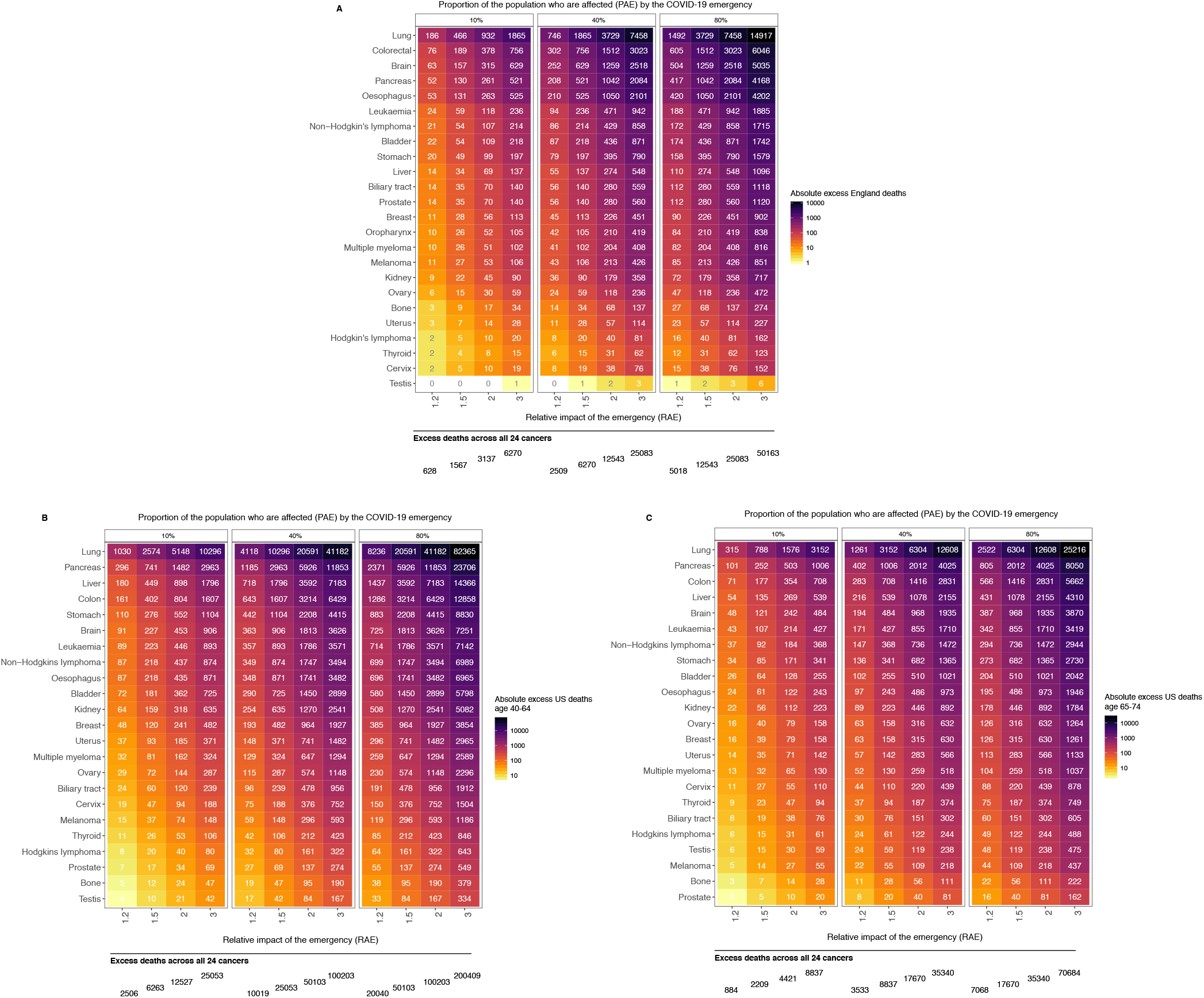
Estimated number of excess deaths at 1 year due to the COVID-19 emergency by cancer site for incident cases (A) scaled up to the population of England aged 30+ consisting of 35 million individuals using England (CALIBER) mortality estimates, (B) scaled up to the population of US aged 40-64 consisting of 208 million individuals using US (SEER) mortality estimates for this age range and (C) scaled up to the population of US aged 65-74 consisting of 61 million individuals using US (SEER) mortality estimates for this age range.

### Estimates of excess COVID-19-related deaths by cancer site for incident cases in US

We found broadly comparable 1-year mortality among incident cancers across cancer sites in England and the US (Figure S2B). For the US, based on SEER incidence and 1-year mortality for individuals aged 40-64, at COVID-19 PAE of 40%, we estimated 10,019, 25,053 and 50,103 excess deaths at RIE of 1.2, 1.5 and 2 respectively, when extrapolating to the US population of this age group(208,284,801 individuals) (Figure 2B). Using mortality estimates for individuals aged 65–74, at COVID-19 PAE of 40%, we estimated 3,533, 8,837 and 17,679 excess deaths at RIE of 1.2, 1.5 and 2 respectively, when extrapolating to the US population of this age group (61,346,445 individuals) (Figure 2C). For a PAE of 40% and RIE of 1.5 (range 1.2-2), we estimated 33,890 excess deaths (range 13,552-67,782) in individuals older than age 40.

### Proportion of 15 comorbidity clusters relevant to COVID-19 risk

Across all cancers, the proportions of 0, 1,2 and 3+ comorbidities were 51.8%, 25.2%, 14.2% and 8.8% respectively for prevalent cancers. Comorbidities were common in people with cancer; e.g., hypertension (34,696 [19.0%]), CVD (23,532 [12.9%]), CKD (9,530 [5.2%]) and obesity (9,491 [5.2%]) (Figure 3). Prevalence of comorbidities in some cancers differed from the condition’s prevalence in the overall population. For example, COPD (3.0%) compared to CVD (12.9%) is a relatively uncommon comorbidity in patients with cancer, except for lung cancer where patients had a higher than background COPD prevalence (25.7%) (Figure 3). Conversely, a ‘metabolic phenotype’ was particularly common in uterine cancer, where a relatively large proportion of patients had hypertension (48.8%), CVD (26.2%), obesity (14.1%) and type-2 diabetes (9.3%) (Figure 3).

**Fig. 3.**
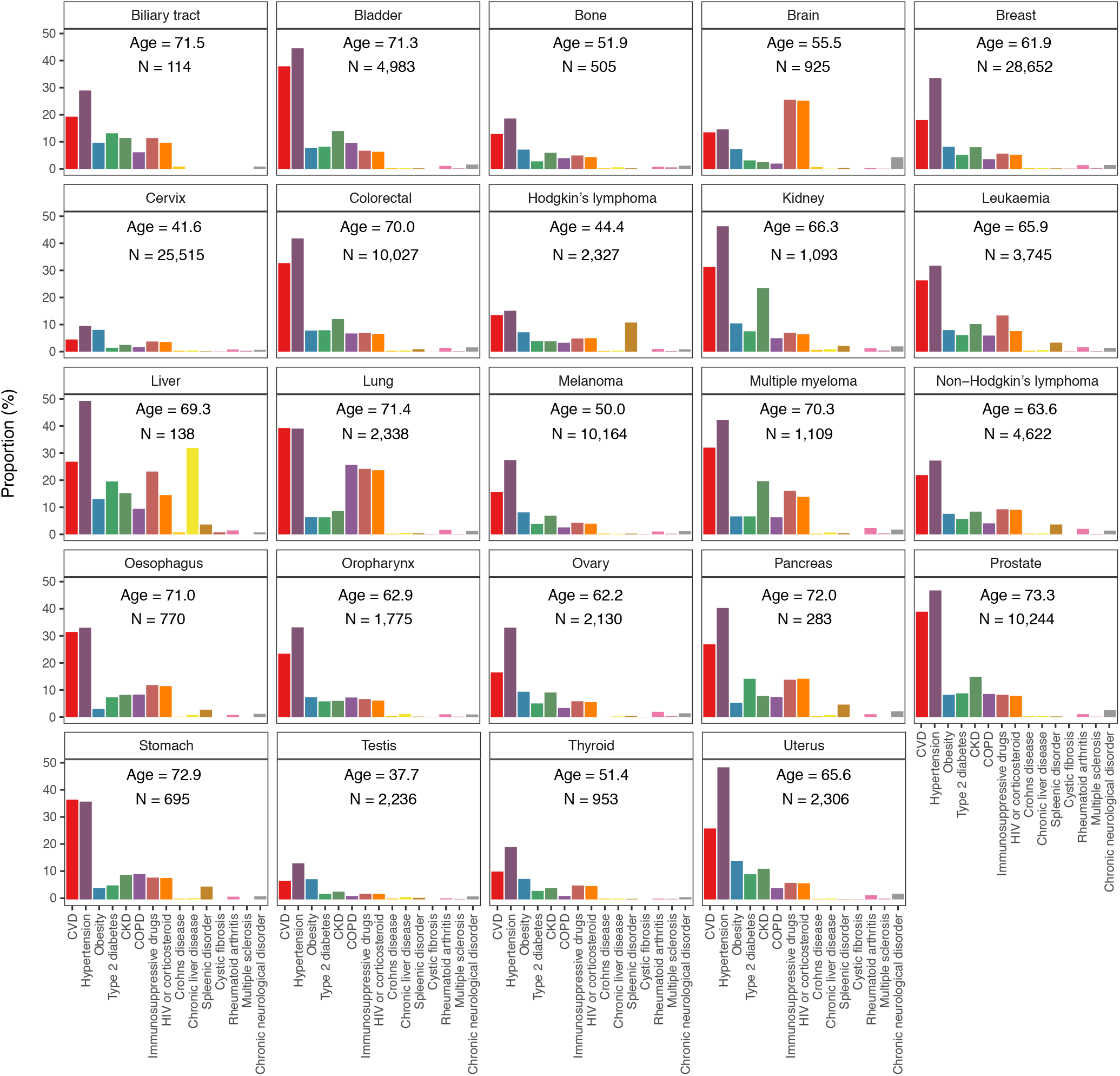
Proportion of patients with any of the 15 comorbidity clusters by cancer site for prevalent cases (N=117,978) in a population of 3,862,012 adults in England (CALIBER). Age indicates mean age at diagnosis.

### Number of comorbid conditions and 1-year mortality in incident cancers

The proportions of individuals with 0, 1, 2 and 3+ comorbidities for incident cancers were 34.8%, 25.6%, 20.5% and 19.2% respectively (Figure 4A). We found in incident cases that multimorbidity (≥3 vs 0 conditions) was associated with a further increase in 1-year mortality. Cancers of the pancreas (65.7% vs. 80.1%), biliary tract (58.6% vs. 64.8%), lung (51.9% vs. 60.9%), brain (46.4% vs. 80.9%) and stomach (42.4% vs. 48.3%) exhibited the most pronounced effects (Figure 4B, Figure S6). Consistent findings were found for prevalent cancer cases (Figure S5, Figure S7)

**Fig. 4.**
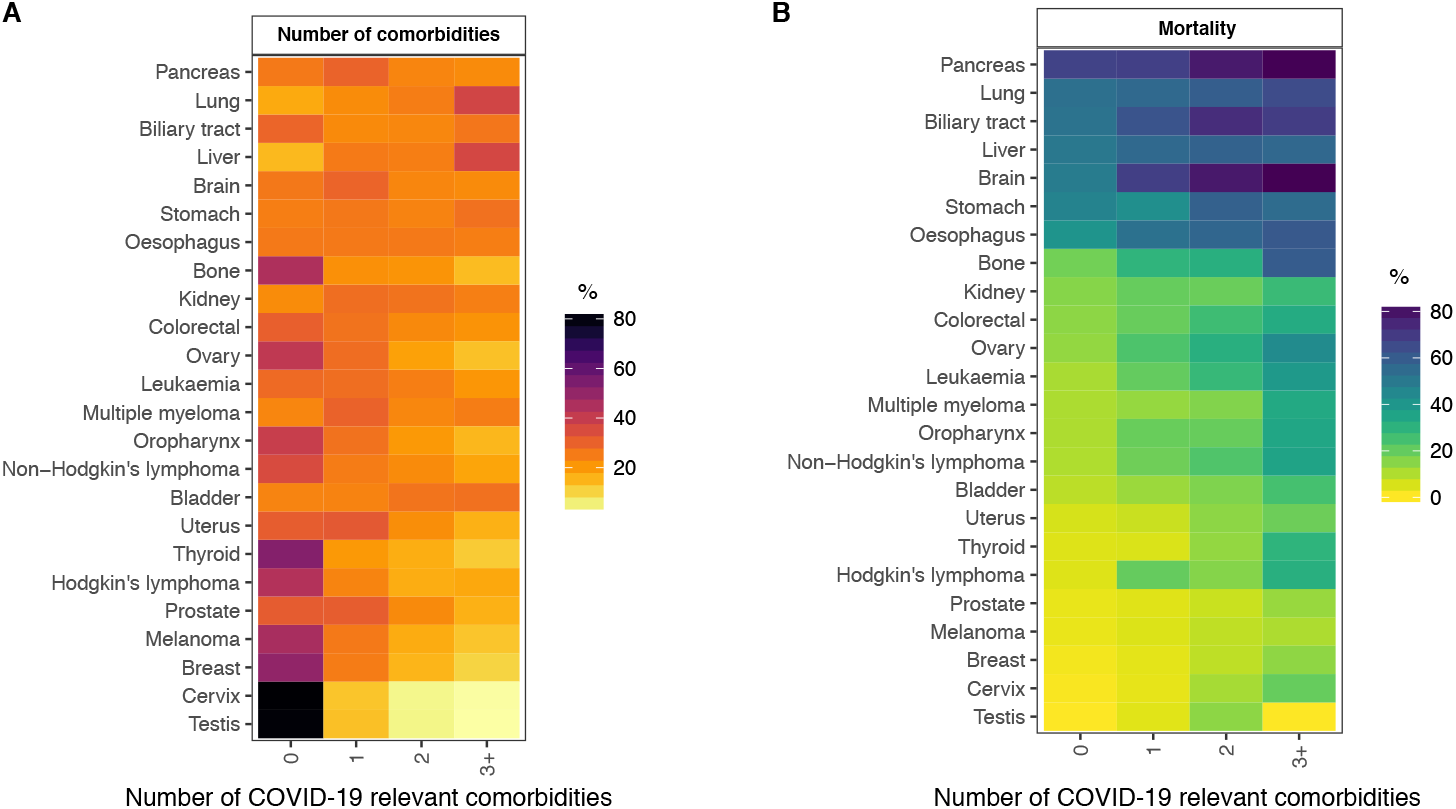
Comorbidity clusters relevant for COVID-19 risk in incident cases in England (CALIBER). (A) Proportion of individuals with 0, 1,2 and 3+ comorbidities by cancer site. (B) 1-year mortality in individuals with 0, 1, 2 and 3+ comorbidities by cancer site.

### Excess 1-year COVID-19-related deaths by cancer site and number of comorbid conditions in England

For incident cancers, when considering COVID-19 PAE of 40% and RIE of 1.5, we observed 1,210,1,509,1,572 and 1,983 excess deaths in individuals with 0, 1, 2 and 3+ non-cancer comorbidities; 5,064 (80.7%) of these deaths occur in patients with ≥1 comorbidities (Figure 5). When considering COVID-19 PAE of 40% and RIE of 1.5 for prevalent cancers, we observed 2,724, 3,480, 2,955 and 2,558 excess deaths in individuals with 0, 1, 2 and 3+ non-cancer comorbidities; 8,993 (76.8%) of these deaths occur in patients with ≥1 comorbidities (Figure S8). When considering both prevalent and incident cancers together at COVID-19 PAE of 40%, we estimated 17,991 excess deaths at RIE of 1.5; 78.1% of these deaths occur in patients with ≥1 comorbidities.

**Fig. 5.**
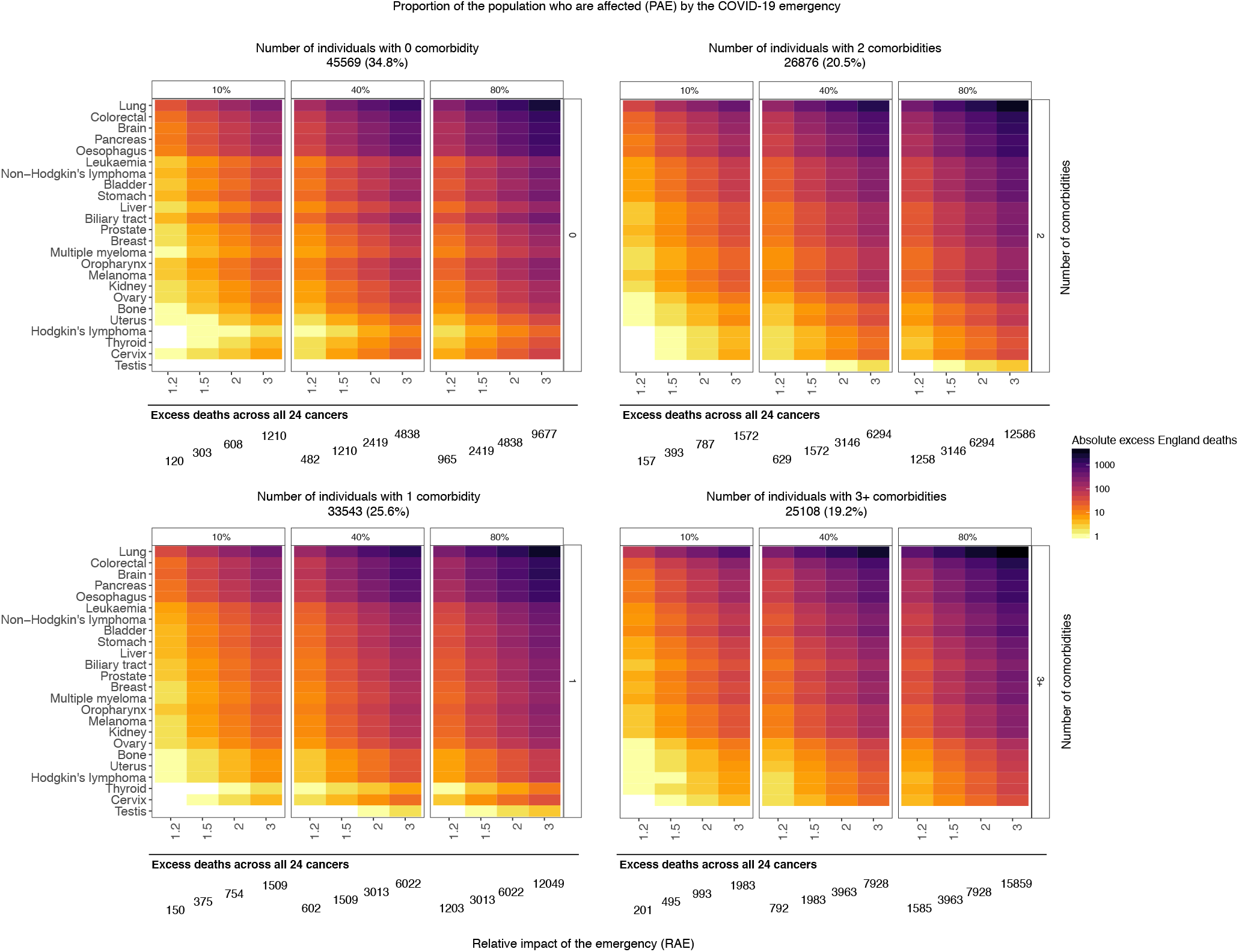
Estimated number of excess deaths at 1 year due to the COVID-19 emergency by cancer site and number of non-cancer comorbidities for incident cases, scaled up to the population of England aged 30+ consisting of 35 million individuals.

## Discussion

This is the first study demonstrating profound recent changes in cancer care delivery in multiple centers and the first study modelling overall excess deaths across incident and prevalent cases of cancer (24 sites) where comorbidities were present in 78.1% of excess deaths.

### Near real-time intelligence

There has been a lack of empirical insights and cancer-specific models to counterbalance infectious disease modeling in informing evolving policy responses to the emergency. We propose that weekly national indicators and warnings for vulnerable patient groups such as cancer patients with multimorbidity are essential. For example, weekly cause-specific mortality reporting of registered deaths and linking of those death records to primary and secondary care records for cancer and other services would allow rapid ‘root cause analysis’ of the extent to which service changes have a net positive or adverse contribution to excess mortality and other non-fatal outcomes.

### Changes in cancer services in response to the emergency

We demonstrated major changes in the delivery of cancer services in the UK, which are also likely to impact on cancer patient survival. Two metrics were chosen to reflect both active treatment for patients diagnosed with cancer and the urgent referral of patients who may have a new diagnosis of cancer. The large declines observed in chemotherapy attendance may reflect workforce/capacity or resources being redirected to care for infected patients (e.g., to intensive care) and the desire of clinicians and patients to minimize the risks of COVID-19 infection(8). We note the importance of considering multimorbidity in chemotherapy prioritization, as the risk associated with COVID-19 increases in patients with multimorbidity. However, some of these patients may benefit from chemotherapy avoidance to mitigate immunosuppression. Additionally, we observed large declines in urgent referrals for early cancer diagnosis. Delays in urgent cancer referrals may be caused by patients struggling to secure appointments due to reprioritized health systems, or patients deciding not to seek care due to perceived risk of COVID-19 infection(17). An unintended consequence of these service changes may be excess deaths via increases in emergency admissions, cancer stage shift at presentation and the undermining of curative/life prolonging surgical resection.

### Modeling impact of the emergency: the ‘untold toll’

We modeled a range of estimates of PAE because the true value is not known. PAE is a summary measure of exposure to the adverse health consequences of the emergency, combining four parameters; the proportion of the population with 1) ill-health in those infected, 2) net adverse health due to changes in health services designed to protect cancer patients from infection, 3) net adverse health consequences of physical distancing, 4) adverse health consequences of economic downturn. Since empirical estimates of each of these four parameters for PAE are not yet available, we chose a PAE range of 10%-80%, with 40% as a plausible estimate.

RIE is a summary measure of the combined impact on mortality of infection, health service change, physical distancing and economic downturn. We modeled a range of estimates of RIE. The only direct evidence we have to date of the magnitude of the RIE comes from national death reporting in England(1). Here, an RIE of 1.5 is consistent with ONS estimates of excess deaths during the COVID-19 emergency in England, which reported almost 8,000 more deaths in the week of 10th April 2020, than the 5-year average of 10,520(1). A higher RIE is possible, given the following evidence: 1) patients with cancer may have twice the odds of developing COVID-19 infection (odds ratio=2.31) compared to the rest of the population(28), 2) COVID-19 causes patients with cancer to deteriorate more rapidly (hazard ratio=3.5) compared to those without cancer(11) and 3) COVID-19 case fatality rates are higher in patients with comorbid conditions including CVD, diabetes, hypertension and cancer(29). For cancer patients of working age, unemployment may affect mortality, as we have previously demonstrated across 75 countries(30). Social isolation is also known to represent a mortality risk in cancer(31, 32). Our findings relate to predicted excess mortality in the next 12 months. However, the socioeconomic effects on health from the current epidemic are likely to last for a considerable period beyond one year(33), meaning our figures are likely a significant underestimate.

### Excess deaths

Based on the available evidence, we estimate an excess of 6,270 excess deaths at 1 year in patients with incident cancers in England. The recorded underlying cause of these excess deaths may be cancer, COVID-19, or comorbidity (such as myocardial infarction), and it is likely that in the COVID-19 emergency, there may be changes in cause-of-death recording. This is why our model uses death from any cause - for which there can be no changes in recording - and which is relevant to multimorbid cancer patients. Our conservative estimate nonetheless represents a significant additional burden of 20%, the total number of cancer deaths annually among incident cases in England is 31,354(34). We present estimates of excess deaths in incident cases, most of whom will be under active surgical, adjuvant or palliative treatments. We also demonstrate excess deaths in those with prevalent cancers, many of whom have survived the initial high-risk period; these data are particularly relevant to ongoing patient management.

### Model application in the US

Using SEER estimates of incidence and 1-year mortality, we modeled excess deaths in the US. For example, with an RIE 1.5 and PAE 40% we estimate 33,890 excess 1-year deaths. Like other cancer registries, SEER does not obtain information on the range of (COVID-19 relevant) comorbidities. We found good agreement between England and US estimates for incidence and mortality(35).

### Importance of multimorbidity

We show that most patients with cancer have non-cancer comorbid conditions which confer additional mortality risk; many of these comorbidities are treatable by non-cancer services. The COVID-19 emergency has prompted new questions about which cancer patients are most vulnerable and how best to mitigate risk. Effective management of hypertension, for example, may avert death and major adverse events, but it is possible that blood pressure control may worsen during the emergency and contribute to excess deaths.

### Policy implications

Our study can inform policy in four areas. First, there is a need to mobilize access to real-time national data on mortality (to determine which disease combinations pose the greatest risk) and on cancer health services activity (to monitor the effects of system change during the emergency on care delivery and future health outcomes). This health services intelligence should include both data from cancer services and from those services (e.g. internists and family physicians) who manage the treatable comorbidities of cancer patients. Second, the triaging of chemotherapy prioritization would usefully incorporate patient-specific risk/benefit assessments to include multimorbidity, particularly in situations where chemotherapy outweighs the benefits of non-treatment and safety issues(17) related to COVID-19. Third, there is a need to enable access to life-saving early diagnosis and to apply a triage system to prioritize urgent referrals for citizens with worrying symptoms, especially those with comorbidities, as multimorbidity may mask an underlying cancer. Fourth, the policy of ‘shielding’ vulnerable patients under active treatment for cancer, or with one of a range of other non-malignant conditions has been implemented in 1.5 million individuals in England, involving home delivery of food parcels and medicines for an initial period of 12 weeks. We propose that estimates of background (pre COVID-19) 1-year mortality risks provides a transparent rationale, readily executable in health records, for identifying priority patient groups including cancer patients with multimorbidities to receive shielding interventions.

### Limitations

This study has important limitations. First, there is a lack of near real-time clinical data in multiple digitally-mature hospitals with large numbers of COVID-19 infected patients. Second, the health records we used may have missed cases of cancer and underestimated incidence(36); if so, our estimates of excess deaths may be conservative. The National Health Service has national linked hospital admissions and cancer registration data with information on stage and details of surgical, chemotherapeutic and radiotherapy treatment of cancer. However, information governance for such data can take months to secure, making data-enabled research and time-sensitive responsive service improvement difficult. Third, we did not have access to multimorbidity data for the US. Fourth, we did not have access to data on children.

## Conclusion

Our data have highlighted how cancer patients with multimorbidity are a particularly at-risk group during the current pandemic. In order to ensure effective cancer policy and avoid excess deaths, both during and after the COVID-19 emergency, it is critical to ensure near-real time reporting of cause-specific excess mortality, urgent cancer referrals and treatment statistics, so as to inform the most optimal delivery of care in this extremely vulnerable group of patients.

## Data Availability

All data are available in the manuscript.

## Authors’ contribution

**Research question:** AL, HH. **Funding:** AL, AB, HH, DATA-CAN. **Study design and analysis plan:** AL, LP, AB, MK, WHC, HH. Preparation of data, including electronic health record phenotyping in the CALIBER **open portal:** AL, LP, SD. **Provision of weekly hospital data:** GH, KPJ, MDF, DH, ML, KB, CD. **Statistical analysis:** AL, LP, MK, WHC. **Drafting initial versions of manuscript:** AL, HH. **Drafting final versions of manuscript:** AL, ML, HH. **Critical review of early and final versions of manuscript:** All authors.

## Acknowledgements

We thank Tony Hagger, Shiva Thapa, Mohammed Emran, Cara Anderson, Louise Herron, Philip Melling and Lee Cogger for their help on collating data on urgent cancer referrals and chemotherapy attendances. We thank Charles Swanton for his valuable comments on the manuscript. This work uses data provided by patients and collected by the NHS as part of their care and support.

## Supplementary methods

This study was performed as part of the CALIBER program (https://caliberresearch.org/portal), which is a research resource consisting of anonymized, coded variables extracted from linked electronic health records, methods, tools and specialized infrastructure. Ethical approval was granted (20_074R2) by the MHRA (UK) Independent Scientific Advisory Committee of the Clinical Practice Research Data Link under Section 251 (NHS Social Care Act 2006). The interpretation and conclusions contained in this study are those of the authors’ alone.

**15 comorbid conditions or condition clusters involving 40 individual conditions defined by the Centers for Disease Control (CDC) or Public Health England (PHE)** as associated risk factors for poor outcomes caused by severe COVID-19 infection. Both lists from CDC and PHE included chronic respiratory disease; chronic heart disease; immunocompromised; HIV or use of corticosteroids; obesity; diabetes; chronic kidney disease; chronic liver disease. The PHE list included additional condition clusters (chronic neurological disorders; splenic disorders). We have performed analyses for all the above conditions and have additionally considered hypertension, Crohn’s disease, cystic fibrosis and rheumatoid arthritis.

Given that condition clusters such as (i) chronic heart disease would involve a range of conditions, we have derived composite variables to include 15 conditions considered as cardiovascular disease (CVD) that included acute myocardial infarction, unstable angina, chronic stable angina, heart failure, cardiac arrest or sudden coronary death, transient ischemic attack, intracerebral hemorrhage, subarachnoid hemorrhage, ischemic stroke, abdominal aortic aneurysm, peripheral arterial disease, atrial fibrillation, congenital heart disease, hypertrophic and dilated cardiomyopathy and valve disease (multiple, mitral and aortic)(23). We also considered (ii) Hypertension, defined as ≥140 mmHg systolic blood pressure (or ≥150 mmHg for people aged ≥60 years without diabetes and chronic kidney disease) and/or ≥90 mmHg diastolic blood pressure(22), (iii) type 2 diabetes, (iv) obesity, defined as a body mass index of ≥30kg/m2, (v) chronic kidney disease (CKD), (vi) chronic obstructive pulmonary disease (COPD)(25), (vii) patients on immunosuppressive drugs (not cancer chemotherapy), (viii) patients with HIV or corticosteroid prescription, (ix) chronic neurological disorders, defined as a composite of Parkinson’s disease, motor neuron disease, learning disability and cerebral palsy, (x) multiple sclerosis separately, (xi) splenic disorders, defined as a composite of splenomegaly, splenectomy and hyposplenism, (xii) chronic liver diseases, defined as a composite of chronic viral hepatitis B or C, primary biliary cholangitis, liver fibrosis, liver cirrhosis and non-alcoholic fatty liver disease, (xiii) Crohn’s disease, (xiv) cystic fibrosis and (xv) rheumatoid arthritis(24).

### Analyses

In CALIBER (England), we estimated the frequency (%) of comorbid conditions in prevalent cancers (diagnosed at or any time prior to baseline) and incident cancers as occurring any time over follow-up. We estimated incidence rates for as the number of new cancers by cancer site per 100,000 person years. We compared CALIBER incidence rates for England with those from the UK from the International Agency for Research on Cancer (IARC). In CALIBER, we generated Kaplan-Meier estimates on 1-year mortality for incident cancers observed from 2012-2016 (by cancer cites and by number of comorbid conditions) and prevalent cancers (by cancer sites and by number of comorbid conditions). We only considered non-fatal incident cases, i.e., alive for at least 30 days following cancer diagnosis to account for potential time lag in death recording. Excess deaths for incident cancers were estimated based on 1-year mortality and 1-year incidence rates, scaled up to a population of 35,407,313 individuals aged 30 and above in England based on population estimates from 2018(37). We used publicly available data from the US from the Surveillance, Epidemiology, and End Results (SEER) program to estimate 1 year mortality as 1 minus the reported survival(27). We considered mortality data from SEER for individuals aged 40-64, 65-74 and 75+. We applied our model (RIE and PAE) to SEER estimates of incidence and 1-year mortality (according to the corresponding age groups) scaled up to the US population of individuals aged 40-64 (208,284,801 individuals) and 65-74 (61,346,445 individuals) based on population estimates from 2018(38).

